# Risk of severe COVID-19 from the Delta and Omicron variants in relation to vaccination status, sex, age and comorbidities – surveillance results from southern Sweden

**DOI:** 10.1101/2022.02.03.22270389

**Authors:** Fredrik Kahn, Carl Bonander, Mahnaz Moghaddassi, Magnus Rasmussen, Ulf Malmqvist, Malin Inghammar, Jonas Björk

## Abstract

The risk of severe COVID-19 disease requiring hospitalization with extensive oxygen supply was compared among infected cases during two calendar periods when Delta and Omicron, respectively, were the dominating virus variants in Scania county, Sweden. Adjustments were made for differences among cases in comorbidities, prior infection, vaccination status, age and sex. Markedly lower risks were observed from Omicron among the vaccinated in the present study. The risk of severe disease was also lower for unvaccinated during Omicron than during Delta, but remained high among older people and middle-aged males with comorbidities. Efforts to increase vaccination uptake across countries, populations and subgroups should thus remain a public health priority.

---

The new SARS-CoV-2 variant of concern (VOC) Omicron (B.1.1.529) was first reported from South Africa on November 24, 2021, and has since then spread rapidly worldwide. Comparisons of secondary attack rates within Danish households have suggested lower protection against infection among vaccinated persons if the index case was Omicron rather than Delta infected (1). Similarly, a Canadian study, utilizing a test-negative design, found that two vaccine doses did not protect against Omicron infection, while a third dose provided some immediate protection but still substantially less than against Delta (2). However, available evidence suggests that the vaccine protection against severe disease from Omicron remains high (3-5). A Danish cohort study among infected individuals found a substantially lower risk of hospitalization with Omicron infection than Delta among both vaccinated and unvaccinated individuals (4). A limitation in this study was the few infections in older age groups, especially among the unvaccinated. How threatening Omicron is to unvaccinated populations, older persons and people with underlying conditions thus merits further investigation. The aim of the present study was to monitor Omicron risks in risk groups defined by sex, age and comorbidities in addition to vaccination status. The study was conducted in Scania, southern Sweden, an ethnically and socioeconomically diverse region exceeding 1.3 million inhabitants and with local variation in vaccination coverage between 40% and 90% among adults.

## Surveillance of COVID-19 cases and vaccination

The study cohort included all persons residing in Skåne, southern Sweden, on December 27^th^, 2020 (baseline) when vaccinations started (n = 1 384 531) (6, 7), and was followed for infection until January 9^th^, 2022 and until January 25^th^, 2022 for hospitalizations and assessment of disease severity. Individuals who died or moved out from the region were censored on the date of death or relocation. The different data sources used in the present study were linked using the personal identification number assigned to all Swedish residents (8). Regional health registers were used for data on comorbidities, defined as diagnoses in inpatient or specialized care at any time point during the last five years before baseline in the following disease groups (Supplementary Table 1): cardiovascular diseases, diabetes or obesity, kidney or liver diseases, respiratory diseases, neurological diseases, cancer or immunosuppressed states, and other conditions and diseases (Down syndrome, HIV, sickle cell anemia, drug addiction, thalasemia or mental health disorder). The number of comorbidities concerning these groupings was counted and classified as zero, one or at least two in the analyses.

Weekly updates on vaccination date, type of vaccine and dose were obtained from the National Vaccination Register, and data on COVID-19 cases (positive SARS-CoV-2 test results) from the electronic system SMINet, both kept at the Public Health Agency of Sweden. The regional health registers were accessed continuously to provide data on positive tests rapidly and to assess disease outcomes. Severe COVID-19 was defined as at least 24 hour-hospitalization five days before until 14 days after a positive test and with a need of oxygen supply ≥ 5 L/min or admittance to an intensive care unit (ICU).

We used data from routine sequencing of samples of infected cases in the Scania region and compared disease risks during three different calendar periods (9): i) Delta dominating VOC, 2021 week 27-47, ii) Transition period, 2021 week 48-51 when Omicron was first observed, and iii) Omicron dominating VOC, 2021 week 52 (74% sample prevalence) and 2022 week 1 (88% sample prevalence).

### Risk of severe COVID-19

A total of 55 269 COVID-19 cases were identified during the three calendar periods of follow-up, of which 437 (0.79%) were classified as severe (Table 1). We used logistic regression (Stata SE 14.2, Stata Corp) to estimate the effect of the transition from Delta to Omicron on the occurrence of severe disease (Supplementary Table 2). The analyses were adjusted for age, sex, comorbidities and prior infection, and additionally for booster dose and time since last dose among the vaccinated. The estimated odds of severe COVID-19 was 40% lower (95% confidence interval 18 – 56% lower) among unvaccinated and 71% lower (95% confidence interval 54 – 82% lower) among vaccinated individuals during the Omicron period than during the Delta period. In Supplementary Table 3 and Figure 1, we present logistic regressions analyses and corresponding estimates of the absolute risks of severe COVID-19 among individuals without prior infection, stratified by age, sex, number of comorbidities, vaccination status (vaccinated, unvaccinated) and period (Delta, Omicron). The risk for severe COVID-19 remained high among unvaccinated, first-time infected, cases of both sexes during the Omicron period in the age group 65+, and also among males in the age group 40-64 years with two or more comorbidities (Figure 1A). The risk of severe COVID-19 among vaccinated cases below 65 years was low for both sexes during Omicron, even in the presence of comorbidities (Figure 1B). Elevated risks for severe COVID-19 remained among vaccinated cases aged 65+ during Omicron only in the presence of at least one (males) or at least two comorbidities (females).

**Table 1.**
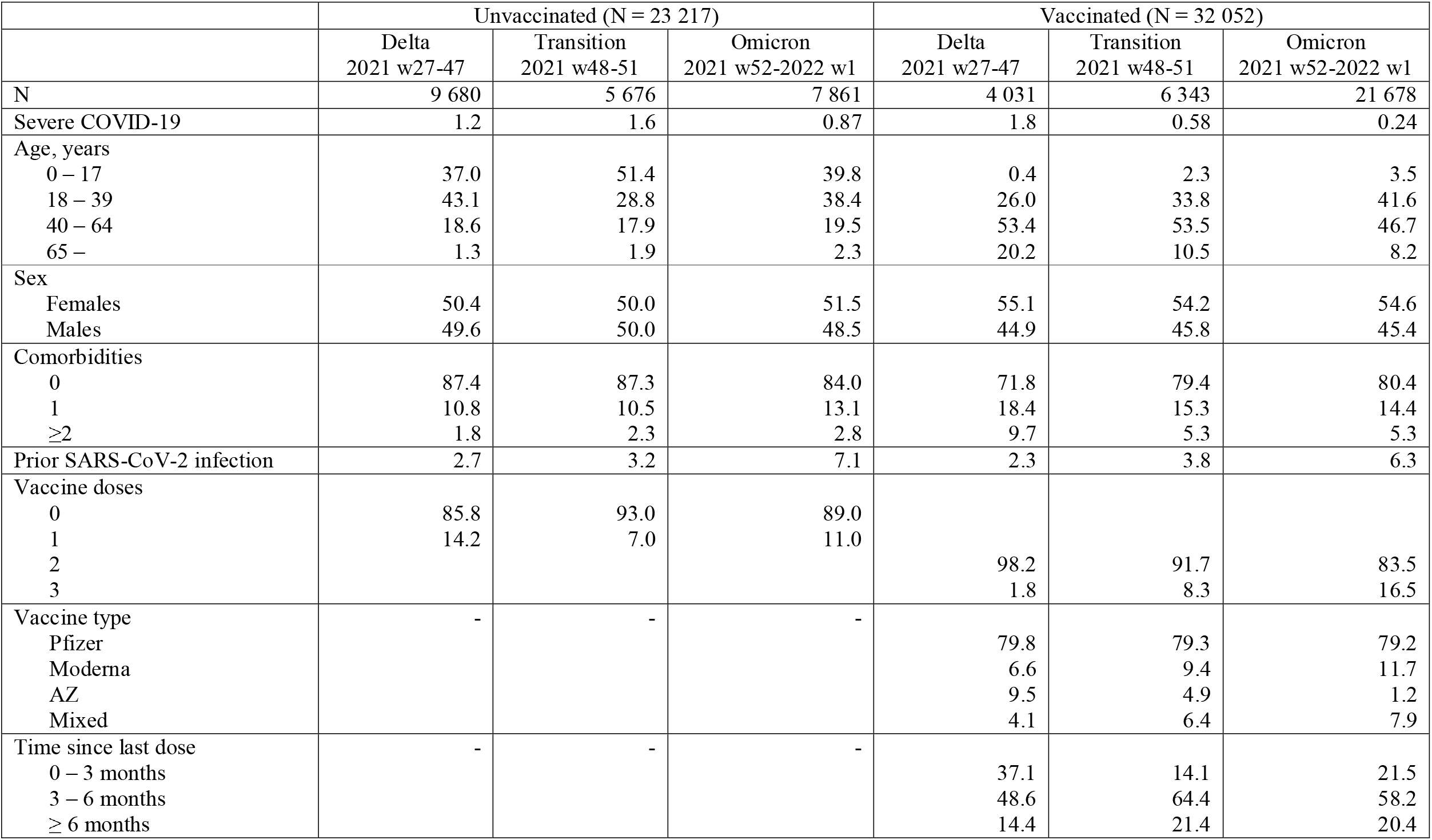
Characteristics (% in each group) of the COVID-19 cases (N = 55 269), stratified by vaccination status and follow up period.

**Figure 1.**
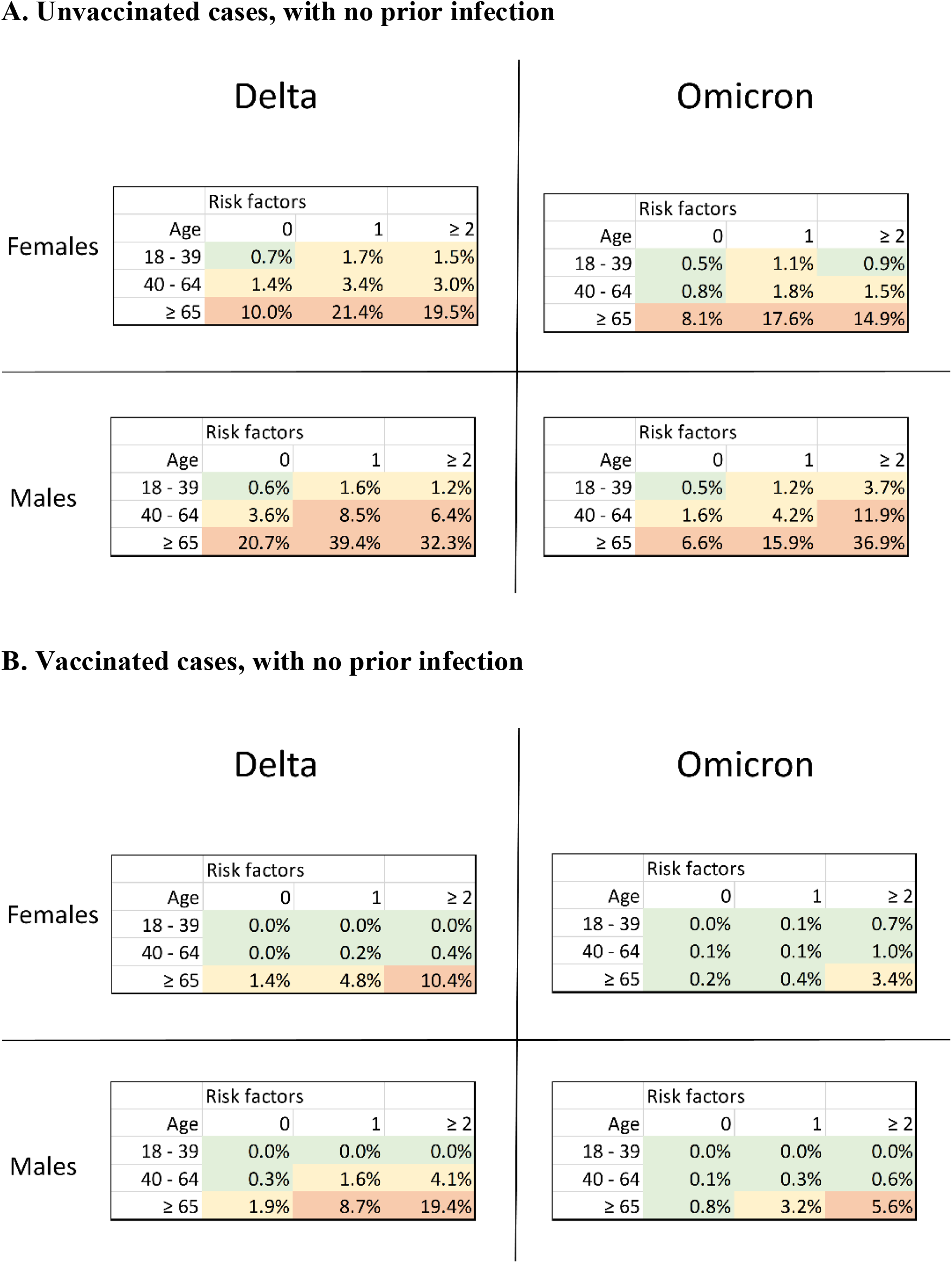
Risks of severe COVID-19 disease among **A**. unvaccinated, **B**. vaccinated cases with no prior infection during the Delta (2021 week 27-47) and Omicron period (2021 weerk 52-2022 week 1), stratified by sex, age and number of risk factors (comorbidities). Green represents risk < 1.0%, yellow 1.0 – 4.9%, orange ≥ 5.0%.

### Vaccine effectiveness

We used continuous density case-control sampling (10) nested within the study cohort together with conditional logistic regression to estimate vaccine effectiveness (VE) against infection and severe COVID-19. A case was defined as a person with a first-time positive test or any positive test at least 90 days after a prior positive test. For each case, 10 controls without a positive test the same week as the case or 90 days prior were randomly selected from the underlying study cohort, matched with respect to sex and age (five-year groups). Median weekly VE against infection was 67% during the Delta period, but showed a declining trend from 2021 week 43 (Figure 2A). A more substantial decline in VE against infection started in the last week (2021 week 51) of the transition period, and by end of follow up when Omicron dominated no vaccine protection against infection remained. VE against severe COVID-19 was estimated monthly, and was stable around 90% across the entire follow up period irrespectively of which VOC that dominated (Figure 2B).

**Figure 2.**
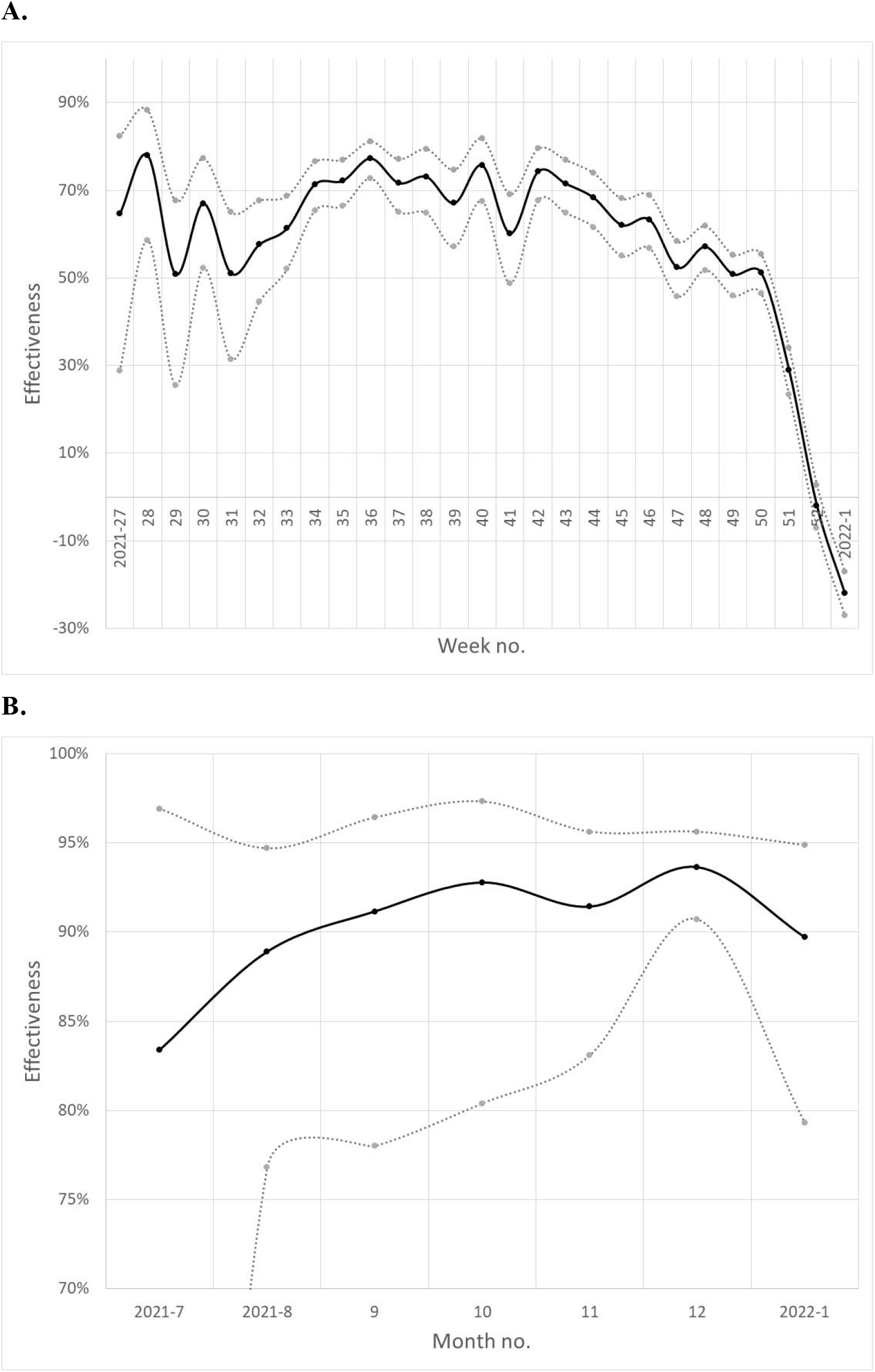
Surveillance in Scania county, southern Sweden, of the estimated vaccine effectiveness after at least two doses against **A**. SARS-CoV-2 infection, weekly 2021 week 27 - 2022 week 1, **B**. severe COVID-19 disease, monthly July 2021 - January 2022, (solid black curves). Grey dotted lines represent 95% confidence intervals.

## Discussion

We observed markedly lower risks of severe disease among the vaccinated during the period when the Omicron VOC dominated. VE thus remained high but changed in nature from protection against both infection and severe disease from Delta to protection only against severe disease from Omicron. The risk of severe disease was also generally lower for unvaccinated persons during the Omicron period, but remained high among older people and middle-aged males with comorbidities. Omicron thus remains a genuine public health concern in countries, populations and subgroups with low vaccination uptake.

The key strengths of our study were the detailed individual-level data on vaccinations, infections and hospitalizations, and the possibility to stratify hospitalized further concerning disease severity, thereby limiting the misclassification of cases hospitalized *with* rather than *because of* COVID-19. A major limitation was that we lacked individual VOC data. This is probably less problematic for the reference period where Delta dominated, but means that some of the cases in the Omicron period were probably infected by Delta. Our estimates may therefore understate the risk reduction associated with Omicron.

## Conclusion

While the Omicron VOC rarely leads to severe disease in vaccinated persons, first-time infection still constitutes a substantial threat to unvaccinated persons with advanced age or underlying illnesses. Efforts to increase vaccination uptake across countries, populations and subgroups should thus remain a public health priority.

## Data Availability

Aggregated surveillance data from the present study will be made available at a web page administrated by the Scania county council.

https://sodrasjukvardsregionen.se/kliniskastudier/covid-vacciner-skyddseffekt/

## Ethics and Permissions

Ethical approval was obtained from the Swedish Ethical Review Authority (2021-00059).

## References

1. Lyngse FP, Mortensen LH, Denwood MJ, Christiansen LE, Møller CH, Skov RL, et al. SARS-CoV-2 Omicron VOC Transmission in Danish Households. medRxiv. 2021:2021.12.27.21268278.

2. Buchan SA, Chung H, Brown KA, Austin PC, Fell DB, Gubbay JB, et al. Effectiveness of COVID-19 vaccines against Omicron or Delta infection. medRxiv. 2022:2021.12.30.21268565.

3. Ulloa AC, Buchan SA, Daneman N, Brown KA. Early estimates of SARS-CoV-2 Omicron variant severity based on a matched cohort study, Ontario, Canada. medRxiv. 2022.

4. Bager P, Wohlfahrt J, Bhatt S, Edslev S, Sieber R, Ingham A, et al. Reduced Risk of Hospitalisation Associated With Infection With SARS-CoV-2 Omicron Relative to Delta: A Danish Cohort Study. SSRN. 2022.

5. Ferguson N, Ghani A, Hinsley W, Volz E. Report 50: Hospitalisation risk for Omicron cases in England.

6. Björk J, Inghammar M, Moghaddassi M, Rasmussen M, Malmqvist U, Kahn F. High level of protection against COVID-19 after two doses of BNT162b2 vaccine in the working age population - first results from a cohort study in Southern Sweden. Infectious diseases (London, England). 2021:1–6.

7. Björk J, Bonander C, Moghaddassi M, Rasmussen M, Malmqvist U, Kahn F, et al. Surveillance of COVID-19 vaccine effectiveness – a real-time case-control study in southern Sweden. medRxiv. 2021.

8. Ludvigsson JF, Otterblad-Olausson P, Pettersson BU, Ekbom A. The Swedish personal identity number: possibilities and pitfalls in healthcare and medical research. European journal of epidemiology. 2009;24(11):659–67.

9. SARS-CoV-2 sequencing in Scania, southern Sweden (in Swedish): Scania County Council; 2022 [Available from: https://www.skane.se/digitala-rapporter/lagesbild-covid-19-i-skane/sekvensering/.

10. Dean NE. Re: “Measurement of vaccine direct effects under the test-negative design”. American journal of epidemiology. 2019;188(4):806–10.

