## Supplementary material for "Risk of severe COVID-19 from the Delta and Omicron variants in relation to vaccination status, sex, age and comorbidities – surveillance results from southern Sweden"

Supplementary Table 1
Supplementary Table 2
Supplementary Table 3A
Supplementary Table 3B

**Supplementary Table 1.** Classification of comorbidities.

| **Disease group** | **ICD-10 codes (incl KVÅ-codes[a])** |
|---|---|
| Cardiovascular diseases | I10-I15, I20-I25, I42-I43<br>I50, I60-I69<br>J81 |
| Diabetes or obesity | E10, E11, E66 |
| Kidney or liver diseases | K70.X, K74.3-K74.6, K75.4, K76.0<br>N18.5, N18.9<br>DR016, DR024 |
| Respiratory diseases | A15-A19<br>E84<br>I26, I27<br>J42, J43, J44, J45, J47, J84<br>J96, J98.2, J98.3 |
| Neurological diseases (including dementia) | G00-G99<br>F00-F03 |
| Cancer or immunosuppressed state (including organ transplantation) | C00-C99<br>KAS, FQA, FQB, JJC, GDG, JLE<br>DR046, DR047, DR048<br>D80.0-D80.1<br>D80.5, D81, D82, D83 |
| Other conditions and diseases<br>• HIV<br>• Thalassemia<br>• Sickle cell<br>• Mood disorders<br>• Schizophrenia spectrum disorders<br>• Substance use disorders<br>• Downs syndrome | B20-B24<br>D56, D57<br>F10-F19, F30-F39, F20-F29<br>Q90 |

[a] Swedish classification of certain interventions during health care visits

**Supplementary Table 2.** Logistic regression analysis for the effect of calendar period on the odds of severe COVID-19 disease, separate among unvaccinated and vaccinated and with adjustment for age, sex, comorbidities, prior infection, time since last dose and booster dose.

| | Unvaccinated N = 23 217 | Vaccinated N = 32 052 |
|---|---|---|
| | OR (95% CI) | OR (95% CI) |
| Background odds | 0.0098 (0.0073 – 0.013) | 0.00084 (0.00029 – 0.0025) |
| Calendar period | | |
| Delta, 2021w27-47 | Ref. | Ref. |
| Transition, 2021 w48-51 | 1.3 (1.0 – 1.8) | 0.58 (0.37 – 0.91) |
| Omicron, 2021 w52 – 2022 w1 | 0.60 (0.44 – 0.82) | 0.29 (0.18 – 0.46) |
| Age, years | | |
| 0 – 17 | 0.11 (0.06 – 0.23) | 0.00[a] |
| 18 – 39 | 1.0 (Ref.) | 1.0 (Ref.) |
| 40 – 64 | 3.5 (2.6 – 4.8) | 3.5 (1.3 – 8.9) |
| ≥ 65 | 20 (14 – 31) | 20 (7.6 – 52) |
| Sex | | |
| Females | 0.58 (0.45 – 0.74) | 0.55 (0.39 – 0.77) |
| Males | Ref. | Ref. |
| Comorbidities | | |
| 0 | Ref. | Ref. |
| 1 | 2.0 (1.4 – 2.7) | 4.8 (2.9 – 8.1) |
| ≥2 | 2.8 (1.8 – 4.2) | 14 (8.3 – 23) |
| Prior SARS-CoV-2 infection | 0.15 (0.04 – 0.62) | 1.3 (0.57 – 3.1) |
| Time since last dose | - | |
| 0 – 3 months | | Ref. |
| 3 – 6 months | | 0.94 (0.51 – 1.7) |
| ≥ 6 months | | 1.3 (0.69 – 2.5) |
| Booster dose | - | 0.84 (0.41 – 1.7) |

[a] No severe cases were observed in this group, and 95% CI could therefore not be calculated

**Supplementary Table 3A.** Logistic regression analysis for the odds of severe COVID-19 disease among unvaccinated persons without prior infection, stratified by sex and calendar period (Delta 2021w27-47 vs. Omicron 2021w52-2022w1).

| | Females, unvaccinated | | Males, unvaccinated | |
|---|---|---|---|---|
| | Delta period OR (95% CI) | Omicron period OR (95% CI) | Delta period OR (95% CI) | Omicr[illegible] OR [illegible] |
| Background odds | 0.0071 (0.0043 – 0.012) | 0.0047 (0.0023 – 0.0099) | 0.0064 (0.0038 – 0.011) | 0.0047 (0.0022 [illegible] |
| Age, years | | | | |
| 0 – 17 | 0.28 (0.096 – 0.85) | 0.00[a] | 0.15 (0.03 – 0.64) | 0.21 (0.[illegible] |
| 18 – 39 | 1.0 (Ref.) | 1.0 (Ref.) | 1.0 (Ref.) | [illegible] |
| 40 – 64 | 2.0 (0.99 – 4.0) | 1.6 (0.59 – 4.5) | 5.8 (3.1 – 11) | 3.5 [illegible] |
| ≥ 65 | 16 (6.3 – 39) | 19 (6.6 – 53) | 41 (18 – 92) | 15 [illegible] |
| Comorbidities | | | | |
| 0 | 1.0 (Ref.) | 1.0 (Ref.) | 1.0 (Ref.) | [illegible] |
| 1 | 2.5 (1.2 – 5.0) | 2.4 (0.96 – 6.2) | 2.5 (1.4 – 4.4) | 2.7 [illegible] |
| ≥ 2 | 2.2 (0.65 – 7.3) | 2.0 (0.62 – 6.5) | 1.8 (0.70 – 4.7) | 8.3 [illegible] |

[a] No severe cases were observed in this group, and 95% CI could therefore not be calculated

**Supplementary Table 3B.** Logistic regression analysis for the odds of severe COVID-19 disease among vaccinated persons without prior infection, stratified by sex and calendar period (Delta 2021w27-47 vs. Omicron 2021w52-2022w1).

| | Females, vaccinated | | Males, vaccinated | |
|---|---|---|---|---|
| | Delta period<br>OR (95% CI) | Omicron period<br>OR (95% CI) | Delta period<br>OR (95% CI) | Omic<br>OF |
| Background odds | 0.00049 (0.000060 – 0.00039 | 0.00058 (0.00020 – 0.0017) | 0.0034 (0.0013 – 0.0090) | 0.00090 (0.00037 |
| Age, years | | | | |
| 0 – 17 | 0.00[a] | 0.00[a] | 0.00[a] | |
| 18 – 39 | 0.00[a] | 0.77 (0.18 – 3.3) | 0.00[a] | |
| 40 – 64 | 1.0 (Ref.) | 1.0 (Ref.) | 1.0 (Ref.) | |
| ≥ 65 | 29 (3.7 – 225) | 3.6 (1.1 – 12) | 5.7 (2.5 – 13) | 9.5 |
| Comorbidities | | | | |
| 0 | 1.0 (Ref.) | 1.0 (Ref.) | 1.0 (Ref.) | |
| 1 | 3.6 (0.88 – 15) | 1.7 (0.32 – 9.0) | 4.9 (1.7 – 14) | 3. |
| ≥ 2 | 8.3 (2.2 – 31) | 17 (4.8 – 58) | 12 (4.5 – 34) | 6.9 |

[a] No severe cases were observed in this group, and 95% CI could therefore not be calculated
